# Barriers and opportunities relating to the implementation and evaluation of a complex digital intervention in England: A case study of Greater Manchester Diabetes My Way

**DOI:** 10.1101/2023.01.27.23285086

**Authors:** Joanna Goldthorpe, Tommy Allen, Martin K Rutter, Joanna Brooks

## Abstract

**Introduction:** Digital diabetes management systems have the potential to deliver cost-effective, flexible self-management support to patients with type 2 diabetes. DiabetesMyWay (DMW) is a platform for an open access website that aims to support diabetes self-management and provide patients with access to their care records. We present a case study focusing on a project involving patients across the large urban area of Greater Manchester (GM).

**Methods:** Interviews were undertaken with 8 members of the project team and 3 patients using the platform. Data were analysed thematically using the template analysis approach.

**Results:** Three themes are presented: Complex approvals, permissions and access across multiple organisations and systems; Capacity challenges in primary care settings; Ongoing support for the project. Implementation and evaluation of GMDMW involved navigating data sharing systems and regulations for complex private and public sector organisations and these impacted on the electronic data sharing necessary for the DMW platform to work as intended. Participants felt that the complexities associated with merging different systems, operated by different organisations, with different data controllers and data owners were greater than anticipated.

**Discussion:** The complexity, number of organisations and levels of governance involved in implementing and evaluating GMDMW were barriers to implementation and evaluation. This reflects inherent problems around mobilising innovation in the NHS. Our findings can help the development and evaluation electronic interventions to improve health to navigate this complex research landscape and ensure that patients can access the most innovative and effective ways to support their health. (245/250 words)

**Author summary:** Digital health interventions and Public Private Partnerships for innovation in the NHS are on the rise and in order to establish efficacy, these need to be evaluated. Often, this research and evaluation is carried out by Universities. However, challenges around navigating governance across these complex, multiple organisations exist where there is a need to share data and collaborate across systems effectively. Here, we describe the many unexpected challenges and solutions to effective data sharing reported by professionals and patients involved in the delivery and evaluation of a UK-based NHS/ private sector collaboration to provide a digital platform to support self-management of diabetes.

## Introduction

DiabetesMyWay (DMW) is a platform for an open access website for people with diabetes that has been available in Scotland since 2008. DMW offers a variety of multimedia resources aimed at improving self-management and from 2010, has offered users access to their clinical data in the form of an electronic personal health record. By 2020 over 60,000 people had registered for DMW (including people with both T2D and type 1 diabetes). Evaluations of the system were encouraging, with 90% of respondents to one survey of users reporting that engagement with DMW helped them make better use of their consultation, improved their diabetes management and improved their condition-related knowledge [1].

Plans for the delivery and evaluation of an enhanced version of DMW across Greater Manchester are described in our original study protocol (2). The project was funded by the National Health Service’s Accelerated Access Collaborative Test Bed Programme. The Test Bed Programme is intended to bring NHS organisations and industry partners together to test combinations of digital technologies in real world settings. The implementation period for GMDMW was punctuated by the Covid-19 pandemic, during which patients with diabetes were considered clinically vulnerable [2,3].

Originally, a census approach to recruitment across the Greater Manchester area was planned; approximately 140k people in the area registered as having T2D were to be offered access to the GMDMW platform from August 2019. 600 were also to be offered additional adjunct digital health interventions: Oviva; Changing Health (both offering behaviour change support) and MyCognition (offering ‘cognitive fitness’ training). However, there have been barriers to implementation and uptake was lower than planned. In 12 April 2022, 628 people had registered to use the platform and 130 of these have the GP enabled permissions to enable all the platform’s functions. By October 2022, this had risen to 4,863 registrations with 370 having GP enabled permissions. Previously, research teams evaluating digital and electronic health innovations have been unable to collect sufficient data on clinical outcomes and have reported barriers to implementation using qualitative methods [4–6]. For this paper, we report lessons learned based on the experiences of stakeholders involved in the implementation and evaluation of GMDMW.

## Methods

Ethical approval was received on 02/09/2019, REC Reference 19/WM/0236, IRAS ID 265621.

### Participants

#### Practitioners/ professional staff

We contacted staff involved in the implementation and delivery of the project via email to invite them to participate. Those interested in taking part contacted the researcher directly to arrange the interview.

Eight staff members involved in implementation and evaluation from relevant organisations took part: NHS (N=4), My Way Digital Health (N=2) and the evaluation team (N=2).

#### Patients

Three T2D patients were recruited via adverts posted on the Diabetes My Way websites. Potential participants were able to contact a member of the research team via email. When contact was made, a participant information sheet was sent to the patient and this was followed up by the researcher.

Given the small sample size, and in order to protect anonymity, individual characteristics of the participants are not reported.

### Interviews

All interviews took place over a video conferencing platform or telephone, according to participant preference and lasted between 27-57 minutes (mean=48 minutes).

### Analysis

An amended template analysis approach was used to structure analysis. Template analysis [7] is a flexible form of thematic analysis which can be adapted to meet the needs of a particular study. It permits the use of a priori themes in advance of coding and analysis to facilitate a focus on specific areas of interest and as a means of guiding initial coding [e.g. King and Brooks, 2017 [7]. The planned process evaluation objectives and exploration of acceptability objectives described in the original study protocol (Goldthorpe et al, in press) were used as an a priori focus for analysis. Analysis was led by JG who undertook manual line by line coding of the data with process and acceptability objectives in mind. This was followed by clustering of initial codes and synthesis of themes in an iterative process until the thematic template developed adequately captured the data. JG and JB met regularly to discuss the evolving analysis with input from the wider research team comprising input from academic researchers, clinicians and clinical network leads. The final template comprised three key themes which are reported here. Quotations are presented verbatim to support findings, with the role and identification number of the participant.

## Results

### ‘It gets complicated pretty quickly’: Approvals, permissions and access across multiple organisations and systems

Difficulties associated with multi-organisation working to deliver the project together with the complexities of merging different systems, operated by different organisations, with different data controllers and data owners were significantly greater than anticipated.

> *If you, sort of, map it all out on a piece of paper the number of interactions between those individuals, it’s quite complex … And each one of those interactions potentially requires a contract in place and a data sharing agreement, and you can imagine, you know, it gets complicated pretty quickly in that sort of situation* (Practitioner/ Professional 1)

The (mostly clinical) team who developed the original application did not anticipate the length of time required to apply, review and receive the necessary approvals from NHS research ethics and research governance required for implementation and evaluation of the project, or the extent of the processes involved.:

> *I think from our point of view we’re obviously NHS based, so the research element of this project has been a bit of an unknown entity, you know, we don’t know some of the processes for ethical approval and, you know, green lights from the university, the HRAs role in it as well*. (Practitioner/ Professional 2)

Obtaining permissions for information governance, data control and data sharing produced unanticipated barriers and delays to implementation and evaluation. For example, the issuing of Patient Identification Centre (PIC) agreements were problematic. These are contracts between NHS sites and the research sponsor: sites that are to be used for the identification and recruitment of patients confirm that they have capacity, plans and necessary governance in place to support the research activity.

During the HRA approvals process, it became apparent that PIC agreements could not be arranged with Clinical Commissioning Groups (CCGs) (formal clusters of General Practices) and rather needed to be authorised with individual practices. This had huge implications on planning for time and resources.

> *Because GPs own the data, they are basically individual components of this big picture. … So, we’ve got, you know, lots of general practitioners that are linked to the NHS, but they’re all individual entities and they don’t have any obligation to share their data with anybody* (Practitioner/ Professional 1)

At the planning stage for DMW in Manchester there had been an assumption that the Scottish pilot study would serve as a roadmap for implementation. However, in practice, a crucial difference existed between the Scottish and English healthcare systems existed in that the Scottish system had patient data sharing agreements for diabetes in place prior to the implementation of DMW.

> *Rather than making the agreements and the connections between the individual providers and Diabetes My Way, they integrated with this data aggregator, which then basically had all the data anyway, so there was no permissions issues, or that sort of stuff. …* (Practitioner/ Professional 3)

In GM the project team dealt with two main groups of NHS organisations with responsibility for the management of data collected and technology used within the NHS: primary care practices and NHS Digital. Adjunct interventions operating from the DMW platform also needed to gain appropriate permissions to interact with patient record systems. General practices control the data for their systems, patients own the data and three separate private companies own the data management platforms. One of the three systems would not engage with the organisations providing the digital platforms for the intervention or the evaluation team (*One of them is really, sort of, not very accommodating to linking up with outside tech or outside interventions* [Practitioner/ Professional 5]), meaning that patients of practices using this system were effectively excluded from GMDMW. In response, the DMW team made a pragmatic decision to initially target practices using one patient record system (EMIS). However, this simplified approach also encountered barriers:

> *So we’ve gone live with the EMIS clinical system first, and that is the vast majority of our GP practices, that’s why we’ve gone with them first. But it’s a process where our technical partner has to speak to EMIS, and EMIS have to kind of go backwards and forwards with NHS Digital*. (Practitioner/ Professional 2)

Researchers experienced multiple barriers to accessing NHS-held patient data for a control group to conduct an analysis of primary and secondary outcomes and cost-effectiveness. Issues were again around data sharing agreements and information governance.

> *Earlier on in the project some of the connections we had were suggesting that we could get the data through various different, sort of, regional paths, through systems that were either developed or in development to do similar things in Greater Manchester. None of those panned out in the end and we ended up getting the data through NHS Digital, through a new dataset that they had compiled* (Practitioner/ Professional 5).

**(N.B**. Accessing data from NHS Digital was delayed due to COVID-19 and the increased demand and pressure this placed on the data processing and applications infrastructure. Ultimately, patient records were accessed via the Greater Manchester Care Record: https://gmwearebettertogether.com/)

### ‘You don’t know how it’s going to work until it’s in the real world’: Implementation challenges in primary care settings

The Covid-19 pandemic and subsequent restrictions in place in the UK since March 2020 meant that engagement with practices was significantly restricted due to extremely limited opportunities to engage with key staff members to overcome identified barriers to data sharing.

> *And then, of course, COVID came along which is another layer of complexity and that meant that, you know, the whole focus of primary care was on other things. (*Practitioner/ Professional 8)
>
> The number of participants accessing the DMW platform has been much lower than planned. This impacted the availability of clinical data required to complete evaluation tasks.

> *We’re going to get even fewer people using it and having two usable blood glucose readings. One, because so few people use the app but two, those that did probably weren’t able to go to their GP practice to get a blood glucose reading done. Or if they were able to, they chose not to because they didn’t want to go to a GP practice* (Practitioner/ Professional 5)

However, whilst Covid-19 may have exacerbated difficulties, broader issues relating to capacity and buy-in existed. To link the GMDMW way platform to patient records required additional activity: before GP surgeries can share information they need permission from individual patients, which requires patients to provide the practice with a formal request to access their data. Once this approval has been registered on the system, digital platforms, with appropriate permissions, can then (theoretically) electronically access personalised patient data based on care records. Once permissions were granted, patients had to enter registration codes to activate the link to their patient records. This process was lengthy and required motivation and action from practice staff and patients to seek and approve the relevant permissions.

> *With the best will in the world, you can test as much as you want, but until it’s real patients trying to use it, that haven’t got the ideal settings set up behind the scenes, then you won’t see some of the unique problems that will come out of a project like this* (Practitioner/ Professional 2)

Given the inherent difficulties involved in registering during the early implementation stage, participants felt that patients who successfully registered and engaged with the intervention were likely to be among the most capable, motivated and persistent. This is supported by the quotes from the patient below:

> *The person that I rang initially wasn’t in, so I had to wait for them to come back in…. Although they’d passed the stuff over to somebody in the GP’s surgery, they didn’t really know what they were doing with it so…and I had some questions and nobody could answer them at the beginning…and then, even when I did speak to the person responsible, she couldn’t answer the questions*. (Patient participant 1)

One participant was concerned that medical language and jargon might cause unnecessary anxiety in patients and felt that this could result in additional workload for primary care staff. They felt that some GPs may feel obligated to make appointments with patients to discuss their records, or edit them before release.

> *I think from April this year [2020] the government has decided that patients who want access to their records should have access to their records. But, like I said before, what they haven’t done is provided the funding along with it to try and discuss the records better with the patients and tell them exactly what they’re seeing, what disease, what values mean and things like that. (*Practitioner/ Professional 7)

A patient we interviewed did indeed report feeling confused in relation to some information accessed via GMDMW. She felt that some information did not relate to her own medical history and that some information had been missed. Although she did not report feeling distressed about this, the information was not helpful to her:

> *I don’t know how it extracts the information from my GPs, but it seemed to have stuff down there that I don’t think I’ve ever had…*. *And you know when you have these sorts of, like quite strange thoughts and you think to yourself, because I do, “has somebody been prescribing stuff in my name for somebody else’s use?”* (Patient participant 2)

### ‘It would yield benefits’: Ongoing support for the GMDMW project

Despite the challenges encountered, significant support remained for the project. Participants viewed the GMDMW platform positively as a potentially useful way to support diabetes self-management in addition to providing additional clinical benefits such as prediction of individual risk:

> *The outcomes for people with diabetes, particularly type 2, across GM are not that good. So, they don’t have, you know, such good control, their management of their risk factors, their cardiovascular receptance is less good, you know, compared to some areas of the country. And the data that I’ve seen as far as My Diabetes My Way is concerned suggested that it would yield benefits in terms of the overall management and the self-management, the self-care that people could achieve* (Practitioner/ Professional 1)

Amongst professionals interviewed there was a perceived need for patients to be able to access an alternative to traditional face to face self-management education sessions, and digital solutions such as DMW were seen as a promising option:

> *The main thing was trying to improve outcomes for the person living with diabetes but additionally the way we were planning to do it was innovative by providing digital solutions and we have a very low uptake of our structured education with regards to diabetes* (Practitioner/ Professional 7)

Participants believed that patients with diabetes were particularly vulnerable to complications associated with Covid-19 and some felt that there were opportunities for DMW to offer support in this context, for example, filling gaps in service provision due to the restrictions around in-person support available at the time of the interview.

> *During COVID and as part of our COVID response that’s going out to practices, we’re saying, “please let your patients know about this [DMW]. We’ve got major issues because of COVID*. (Practitioner/ Professional 8)

Clinicians felt that digital interventions were likely to continue playing a major part in diabetes management beyond the pandemic and that the DMW platform could play a role in this change in delivery.

> *So, our argument is that, you know, in the COVID pandemic people have had less access to primary care and therefore those people that have taken up the intervention, Diabetes My Way potentially benefit more because of that action that they’ve taken* (Practitioner/ Professional 1)

Patients reported that the DMW platform delivered credible information. They valued the ability to access their diabetes-related medical information and have it presented visually. The graphs and “dartboard” visuals on the Diabetes My Way platform were cited as being particularly useful:

> *So, for example, since I’ve had this, I’ve been looking at the HbA1C results, and there seems to be a bit of a trend in terms of, it always seems to get worse over the winter period…*.*So, perhaps being aware of that, and trying to be even more diligent with exercise and diet, to see if that makes any difference (*P1)

Practitioners similarly felt that they could trust the information on the DMW platform to be trustworthy and evidence-based. Access to personalised data available through the DMW platform was perceived as supporting patients to get the most out of their face-to-face consultations, as well as supporting self-management, and knowledge about their own condition:

> *Initial feedback from the people who took up the thing, they found that, yes, it was very useful…ideally what we were thinking is to empower the person with diabetes to manage their own condition better… so they can make informed choices about what they do with their diet, exercise et cetera, and also enable better consultations with the clinicians as well* (Practitioner/ Professional 7)

## Discussion

We developed three themes from the data: ‘It gets complicated pretty quickly’: Approvals, permissions and access across multiple organisations and systems; ‘You don’t know how it’s going to work until it’s in the real world’: Implementation challenges in primary care settings; ‘It would yield benefits’: Ongoing support for the GMDMW project. Participants reported barriers around implementation of the GMDMW system that meant patients were unable to access the self-management interventions in the numbers expected and we could not carry out the evaluation study as originally planned. However, we have captured some valuable learning from the experience, which can help planning for implementation and evaluation of future electronic health innovations.

The diversity of organisations and individuals involved in enabling GMDMW was extremely complex. Patients generate (via attendance at GP practices) and engage with (via GMDMW platform) their own data to self-manage their diabetes. For this to happen, GP practices collect data inputted into electronic patient record systems during consultations and enable sharing after patients have given permission. Private sector companies own and manage the systems that hold the data. NHS Digital collate and manage large amounts of data from these systems for health care intelligence. Finally, for the evaluation of this system to take place, the research team, employed by a University, needed to access this data.

These complex systems need to be co-ordinated and relevant NHS governance and approvals put into place. For implementation of GMDMW, this included HRA approval for recruitment of patients, sign off on Participant Identification Centre agreements for primary care practices to agree to participation including obtaining patient’s permission to share their data with GMDMW. For the evaluation to succeed, approvals from NHS Digital around access to control data were also needed. Insufficient time and resources were available to enable the data linkage and governance needed to complete the implementation and evaluation of GMDMW in the 24 months originally allocated.

The website of the NHS Health Research Authority (HRA) (“HRA Approval – one year on - Health Research Authority,”) [8] states that in 2017 the median time from application via the Integrated Research Application System (IRAS) to approval being granted was 60 days.However for complex cases this can take as long as 18 months [9]. Health and care researchers and organisations who wish to implement and evaluate complex interventions within the NHS should consider the potential barriers to accessing and linking datasets, paying particular care to the multiple layers of permissions and governance that need to be obtained and the length of time needed to negotiate relevant systems.

Although the COVID-19 pandemic affected engagement with Primary Care services and subsequently slowed down activation of the permissions GMDMW required to access patient data, opportunities for research arose based on the mobilisation of data and digital responses to support pandemic management and intelligence [10]. The research team were able to access suitable control data from the Greater Manchester Care Record which was facilitated via use of the Control of patient information (COPI) notice which allowed for the sharing of patient data for COVID-19 purposes [11].

A general practitioner who participated in the research was concerned that extra resources would be needed to support patients to understand and interpret healthcare data, citing potential for misunderstandings and the generation of anxiety, particularly around medical jargon and interpretation of test results. This concern was supported by reports from a patient who felt that some entries in her records might be erroneous. However, research carried out in Sweden and the USA suggests that, despite initial concerns, patients are able to access and understand their health records without over-burdening care providers [12,13]. Further research is needed to explore whether this is the case for diabetes patients in a UK context.

Despite the barriers to implementing and evaluating GMDMW identified by this study, the three patients who were able to access the platform did find it acceptable. They liked the way the platform translated and presented their data with user-friendly graphics, and practitioners feel that DMW could be a valuable support tool for patients with diabetes. This only further highlights the need for solutions to the issues of governance and data linkage reported in this paper to be found, so that researchers can successfully test innovations and patients can ultimately benefit from the latest developments in healthcare.

In conclusion, the complexity, number of organisations and levels of governance involved in implementing and evaluating GMDMW caused the barriers to implementation and evaluation we found in this study. This reflects inherent problems around mobilising innovation in Public-Private Innovation Partnerships (PPIPs) reported by Hammond et al., [4] in their case study of implementation of another NHS Test Bed. The authors cite “magical thinking” around the axiomatic virtue of innovation as leading to a failure of rationalisation and planning [4].

We do however hope that our findings can help those involved in the development and evaluation electronic interventions to improve health to navigate this complex research landscape and ensure that patients can access the most innovative and effective ways to support their health.

## Data Availability

For reasons of confidentiality and risk of identification, the data will not be placed in a public repository. However, selected extracts from the text can be made available on request from the corresponding author.

## Acknowledgements

The authors would like to thank Martina Ratto (MSc) from the Beingwell Group and Debbie Wake, CEO and Chief Medical Officer for My Way Digital Health for fact checking and providing comments regarding some of the technical aspects of implementation.

